# Oral anticoagulation in heart failure complicated by atrial fibrillation: Outcomes in routine data

**DOI:** 10.1101/2023.05.02.23289428

**Authors:** Martin Möckel, Samipa Pudasaini, Henning Thomas Baberg, Benny Levenson, Jürgen Malzahn, Thomas Mansky, Guido Michels, Christian Günster, Elke Jeschke

## Abstract

**Background:** Aim of this study was to test if oral anticoagulant (OAC) use in patients with heart failure (HF), accompanied by atrial fibrillation (AF), leads to a favorable outcome. Further, the specific impact of non-vitamin K oral anticoagulants (NOACs) is analyzed.

**Methods:** Anonymous data from all patients with a health insurance at the largest entity which covers approximately 30% of the German population. Patients with a claims record for hospitalization with the main diagnosis of HF and AF from the years 2017 to 2019 were included. A hospital stay in the previous year was an exclusion criterion. Mortality and readmission for all-cause and stroke/intracranial bleeding (ICB) were analyzed 91-365 days after the index hospitalization. Kaplan-Meier survival curves and multivariable Cox regression models were used to evaluate the impact of medication on outcome.

**Results:** 180,316 cases were included [81 years (IQR 76 to 86), 55.6% female, CHA_2_DS_2_-VASc score ≥ 2 (96.81%)]. In 80.6%, OACs were prescribed (vitamin K antagonists (VKA): 21.7%; direct factor Xa inhibitors (FXaI): 60.0%; direct thrombin inhibitors (DTI): 3.4%; with multiple prescriptions per patient included). The mortality rate was 19.1%, readmission rate was 29.9% and stroke/ICB occurred in 1.9%. Risk of death was lower with the any OAC (HR 0.77, 95% CI 0.75 to 0.79) but without significant differences in OAC type (VKA: HR 0.73, 95% CI 0.71 to 0.76; FXaI: HR 0.77, 95% CI 0.75 to 0.78; DTI: HR 0.71, 95% CI 0.66 to 0.77). The total readmission rate (HR 0.97, 95% CI 0.94 to 0.99) and readmission for stroke/ICB (HR 0.71, 95% CI 0.65 to 0.77) was lower with OAC.

**Conclusions:** Routine data confirm the positive effect of OACs in HF-AF. There are no additional benefits regarding mortality with the use of NOACs.

**Clinical Perspective:** *What is new?:*

- This routine data analysis on a study population of 180,316 indicates a decreasing mortality rate, 91 to 365 days after index hospital stay, in patients with HF complicated by AF in case oral anticoagulants (OACs) were prescribed.
- Secondly, the findings imply no additional benefits of new OACs (NOACs) compared to vitamin K antagonists.

*What are the clinical implications?:*

- Our study highlights the benefits of a permanent oral anticoagulation therapy in patients with heart failure (HF) complicated by atrial fibrillation (AF).
- For patients with HF and AF, the results indicate room for personalizationin choosing the specific OAC type for anticoagulation as NOACs show no survival benefit over vitamin K antagonists.

## Introduction

In developed countries, 1-3% of the population suffers from heart failure (HF) with a presumably high number of additional cases that are yet undiagnosed ^1, 2^. These patients can be severely burdened with their symptomatic complaints and face a reduced health-related quality of life as well as an increased mortality risk ^2–4^. Frequently, HF occurs in synopsis with atrial fibrillation (AF). In this combination, in the following being named as HF-AF, both diseases tend to accelerate each other and thus worsen the general prognosis ^5^. Due to the rising life expectancy, the overall sum of patients presenting with HF-AF is rapidly increasing as age is considered a relevant risk factor for both ^2, 6^. The occurrence of AF in HF patients is currently estimated at one third, varying by HF subtype ^6, 7^. Both constitute to the primary causes for hospitalization ^8^.

Adhering to therapeutic guidelines can decelerate the progression of HF accompanied by AF, e.g. by breaking the vicious cycle of readmissions and increasing mortality risk, and therefore relieving patients and the health care system. In case HF and permanent/persistent/paroxysmal AF co-exist, the *European Society of Cardiology* (ESC) and the American College of Cardiology (ACC)/ American Heart Association (AHA) currently recommends a permanent oral anticoagulation ^9, 10^. This is meant to prevent thromboembolic complications which HF patients are strongly prone to, based on an upregulation of thrombin-activated pathways ^11^. Concomitant AF further elevates this risk ^5, 12^. To address this state of hypercoagulation, a strong indication is applicable for male HF-AF patients with a CHA_2_DS_2_-VASc score of ≥ 2 and for women with a score of ≥ 3. Hereby, new oral anticoagulants (NOACs) are preferred, except for cases of valvular AF in which Vitamin K antagonists (VKAs) represent the gold standard. If CHA_2_DS_2_-VASc scores are at 1 in men or 2 in women, oral anticoagulant (OAC) therapy in HF-AF patients can be individually considered ^9, 13^. In patients with a HF in sinus rhythm, however, OAC use is not indicated for routine use due to the risk of bleeding ^14^. Despite these clear recommendations and the extremely high numbers of patients affected by this disease combination, there is lack of long-term data on the outcome of a treatment with OACs ^5, 15^. Existing studies primarily focus on AF and the use of VKAs or NOACs in this patient group, while effects of OACs in patients with a simultaneous HF were only investigated as part of a smaller, additional subgroup analyses. Therefore, they were based on relatively low sample sizes of < 10,000 AF-HF participants each ^15–19^. The respective randomized controlled trials (RCTs) were pooled regarding their AF-HF sub-cohort, in a meta-analysis by Xiong et al., who reported superiority of single-/high dose NOAC use compared to warfarin, a VKA type, regarding stroke/systemic embolism and occurrence of major bleeding. The authors, however, did not draw conclusions on whether OAC use generally leads to a favorable outcome compared to a non-OAC HF-AF cohort ^20^. The same applies to a meta-analysis by Savarese et al., which confirmed the thesis of additional NOAC benefits in comparison to warfarin ^21^. In a further study, Lip et al. assessed the composite one-year outcome of stroke/thromboembolism/transient ischemic attack/death in AF patients with/without HF and reported a high rate of mortality despite an OAC use in 82.1% of the AF-HF cases ^22^. Referring to the existing literature, a study setting of HF-AF patients with OAC therapy versus non-OAC use is not yet provided. This, however, would be necessary to examine mortality and morbidity outcomes after OAC use in the outline before performing ancillary analysis on OAC subtypes.

By analyzing a nationwide routine data set, this study assesses the outcome of patients suffering from HF and concomitant AF who were treated with OACs after their index hospital stay. The primary hypothesis was the assumption that the use of OACs resulted in a favorable outcome regarding the mortality rate 91-365 days after discharge. Further clinically relevant outcomes e.g., all-cause readmissions as well as the subgroup of hospitalizations resulting from the occurrence of stroke or intracerebral bleeding (ICB) were assessed.

## Methods

### Study design and Study population

For this observational study, pseudonymized nationwide administrative claims data of the *Allgemeine Ortskrankenkasse* (AOK) from the time span between 1^st^ January 2017 and 31^st^ December 2019 were used. The AOK provides health care insurance for approximately 30 percent of the German population and is the largest nation-wide provider of statutory health care insurance in Germany ^23^. Billing data for inpatient treatment, including diagnoses, as well as patient data with information on age, sex, insurance status (i.e. continued/terminated AOK membership) and survival were evaluated. Diagnoses were encoded according to the International Classification of Diseases (ICD-10). Healthcare and health insurance providers jointly issue binding guidelines for coding of diagnoses and procedures in German hospitals. Hospital billing data are thoroughly checked by the Medical Review Board of the Health Insurance Funds and are returned to hospitals for correction if necessary.

Patients with main diagnosis of HF (I11.0, I13.0, I13.2, I50) and diagnosis of AF (I48) were included. Exclusion diagnoses were congenital anomaly (Q20-Q28), transplantation (T86, Z09.80, Z94.1, Z94.3), asthma (J45, J46), and age < 30 years. Further, admission due to HF in the previous year and incomplete medication data within 90 days after the index hospital stay were reasons for exclusion.

### Exposure

Medication was assessed by claims data. Drugs are routinely encoded to the WHO ATC/DDD Index (German modification). OAC was defined as at least one prescription 90 days before the index hospital stay or at least one prescription within 90 days after the hospitalization period with the codes B01AA (VKA), B01AE (Direct thrombin inhibitor: dabigatran, DTI) or B01AF (Direct factor Xa inhibitors: rivaroxaban, apixaban and edoxaban, FXal) according to the Anatomical Therapeutic Chemical Classification.

### Outcomes

The primary outcome was all-cause mortality within 91-365 days after discharge. Further endpoints were readmission for any cause and rehospitalization for ischemic stroke (I63, I64) or ICB (I61) 91-365 days after discharge, respectively.

### Statistical analysis

Descriptive statistics including medians, inter-quartile ranges (IQRs), and proportions were used to describe the study sample. Baseline patient characteristics were compared using Pearson’s Chi-squared test for categorical variables. Subgroup analyses were performed for type of OAC (VKA, FXal, DTI).

Unadjusted survival was estimated using the Kaplan-Meier method, with stratification by OAC. The log-rank test was applied here. Multivariable Cox proportional hazards models were estimated to evaluate the effect of OACs on the different endpoints while adjusting for patient risk factors. Cluster-robust standard errors were applied to account for clustering of patients in hospitals. Models were adjusted for patient age, sex, *New York Heart Association* (NYHA) stage, coronary heart disease, aortic valve disorder, mitral valve disorder, dilated cardiomyopathy, prior ischemic stroke or ICB, ventricular tachycardia, chronic obstructive pulmonary disease (COPD), pneumonia, dementia, Elixhauser comorbidities ^24^, and year of admission. Elixhauser condition cardiac arrhythmia was split into atrial fibrillation and other cardiac arrhythmia. Only those conditions with p < 0.1 on prior univariate analysis were included in the multivariable models. Comorbidities were entered as separate dichotomous variables. Patient age was entered as a continuous variable. Adjusted hazard ratios (HR) and 95% confidence intervals (CI) were calculated. The proportional hazards assumption was evaluated by plotting scaled Schoenfeld residuals against time. All analyses were performed using STATA 16.0 (StataCorp LP, College Station, Texas).

### Ethical approval

This work was performed by using anonymous health insurance data. The principles for good clinical practice and data protection were fully applied and were in accordance with institutional guidelines. This type of study design does not require an ethical approval.

### Data access and availability

Data sharing is not applicable to this article as claim data were analyzed. M.M. and E.J. had full access to all the data and take responsibility for its integrity and the data analysis.

## Results

### Study Cohort

A total of 180,316 patients fulfilled the inclusion criteria (Figure (Fig.) 1). Subdivided by year, 60,150 (2017), 58,054 (2018) and 62,112 (2019) patients were included. The median age in the whole study group was 81 years (IQR, 76 to 86 years). Female patients accounted for 55.6%. The majority of the patients had a CHA_2_DS_2_-VASc score ≥ 2 (96.81%). Most observed comorbidities in the whole study cohort were hypertension (81.8%), renal failure (51.0%) and coronary heart disease (41.8%). 42.6% and 40.4% of the cohort were classified as NYHA stage III or IV, respectively. Table (Tab.) 1 provides a detailed overview of patient characteristics.

**Figure 1.**
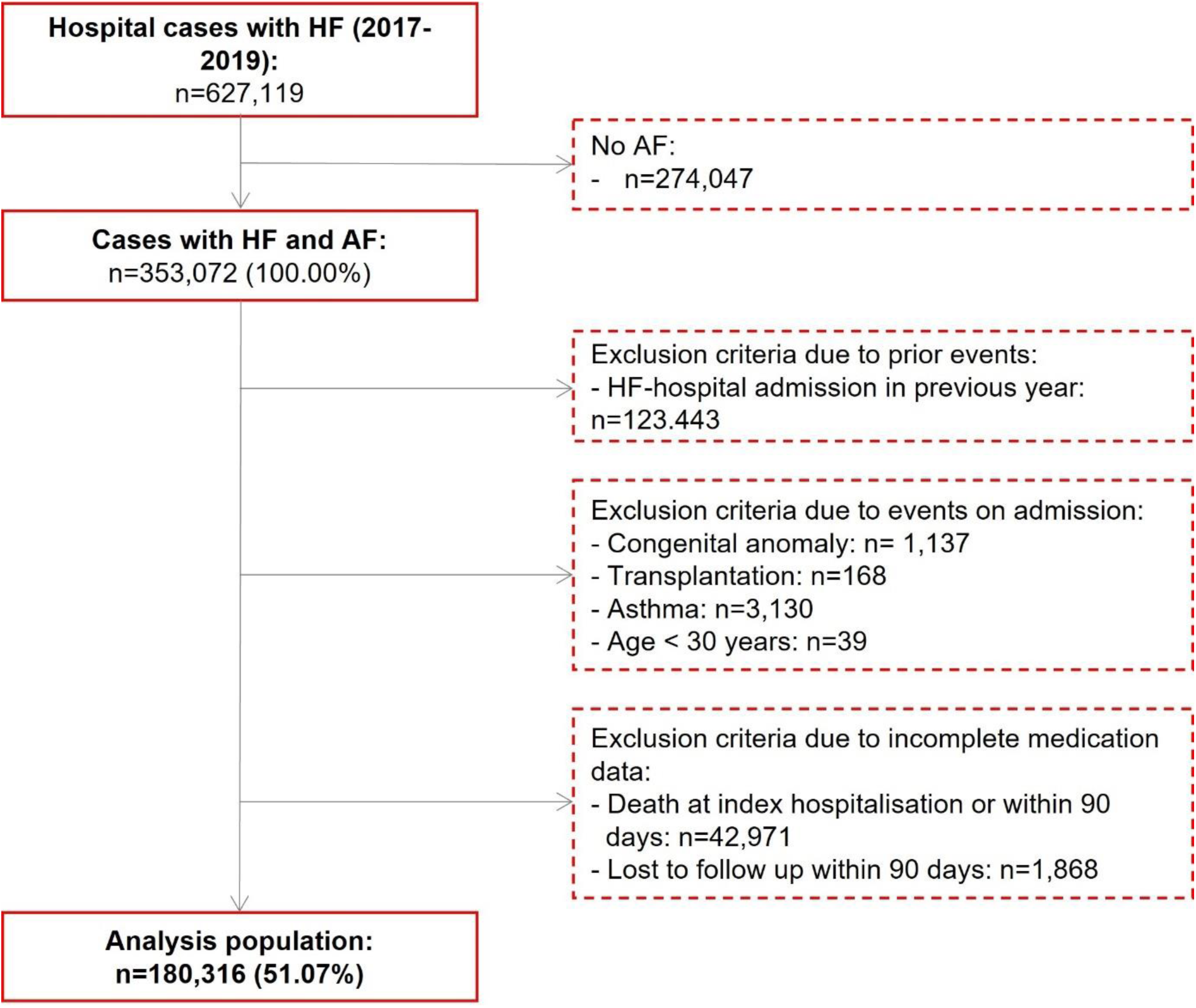
Selection criteria of the study population. Patient flow diagram showing the inclusion and exclusion criteria leading to this study’s analysis population of 180,316 out of 627,119 hospital cases of heart failure between 2017-2019. Exclusion criteria were HF without a co-occurrence of AF, hospital stay in the previous year as well as congenital anomalies, transplantation or asthma as comorbidities. Further, patients aged under 30 years were likewise excluded. Abbreviations: HF heart failure; AF atrial fibrillation.

**Table 1.**
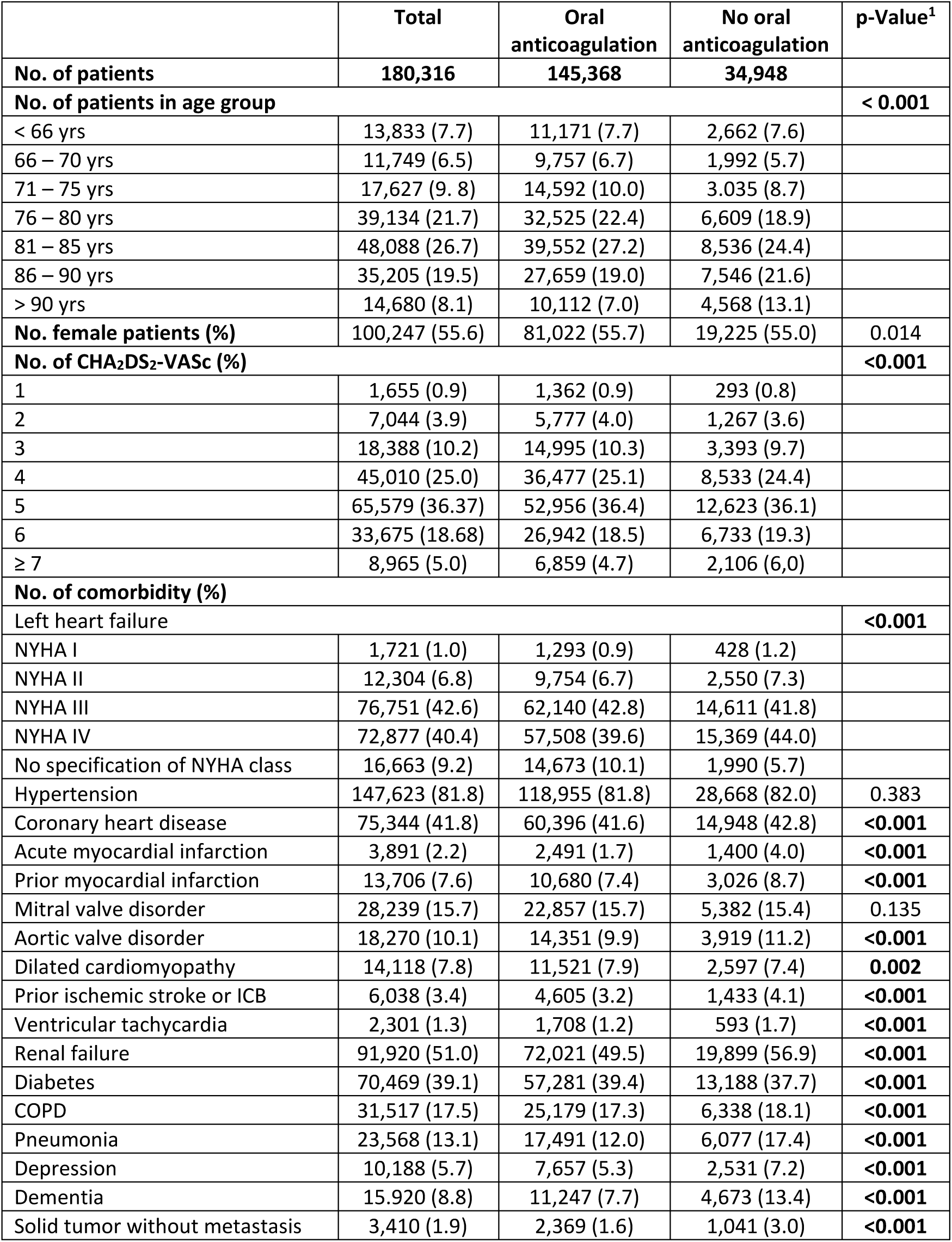
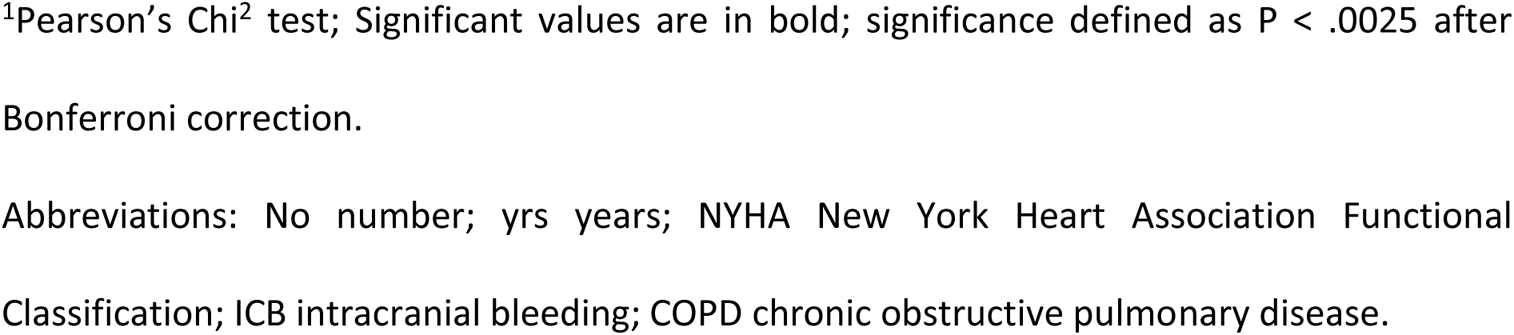
Basic description of patients according to oral anticoagulation.

|  | Total | Oral<br>anticoagulation | No oral<br>anticoagulation | p-Value<sup>1</sup> |
| --- | --- | --- | --- | --- |
| **No. of patients** | **180,316** | **145,368** | **34,948** |  |
| **No. of patients in age group** |  |  |  | **< 0.001** |
| < 66 yrs | 13,833 (7.7) | 11,171 (7.7) | 2,662 (7.6) |  |
| 66 – 70 yrs | 11,749 (6.5) | 9,757 (6.7) | 1,992 (5.7) |  |
| 71 – 75 yrs | 17,627 (9.8) | 14,592 (10.0) | 3,035 (8.7) |  |
| 76 – 80 yrs | 39,134 (21.7) | 32,525 (22.4) | 6,609 (18.9) |  |
| 81 – 85 yrs | 48,088 (26.7) | 39,552 (27.2) | 8,536 (24.4) |  |
| 86 – 90 yrs | 35,205 (19.5) | 27,659 (19.0) | 7,546 (21.6) |  |
| > 90 yrs | 14,680 (8.1) | 10,112 (7.0) | 4,568 (13.1) |  |
| **No. female patients (%)** | 100,247 (55.6) | 81,022 (55.7) | 19,225 (55.0) | 0.014 |
| **No. of CHA<sub>2</sub>DS<sub>2</sub>-VASc (%)** |  |  |  | **<0.001** |
| 1 | 1,655 (0.9) | 1,362 (0.9) | 293 (0.8) |  |
| 2 | 7,044 (3.9) | 5,777 (4.0) | 1,267 (3.6) |  |
| 3 | 18,388 (10.2) | 14,995 (10.3) | 3,393 (9.7) |  |
| 4 | 45,010 (25.0) | 36,477 (25.1) | 8,533 (24.4) |  |
| 5 | 65,579 (36.37) | 52,956 (36.4) | 12,623 (36.1) |  |
| 6 | 33,675 (18.68) | 26,942 (18.5) | 6,733 (19.3) |  |
| ≥ 7 | 8,965 (5.0) | 6,859 (4.7) | 2,106 (6.0) |  |
| **No. of comorbidity (%)** |  |  |  |  |
| Left heart failure |  |  |  | **<0.001** |
| NYHA I | 1,721 (1.0) | 1,293 (0.9) | 428 (1.2) |  |
| NYHA II | 12,304 (6.8) | 9,754 (6.7) | 2,550 (7.3) |  |
| NYHA III | 76,751 (42.6) | 62,140 (42.8) | 14,611 (41.8) |  |
| NYHA IV | 72,877 (40.4) | 57,508 (39.6) | 15,369 (44.0) |  |
| No specification of NYHA class | 16,663 (9.2) | 14,673 (10.1) | 1,990 (5.7) |  |
| Hypertension | 147,623 (81.8) | 118,955 (81.8) | 28,668 (82.0) | 0.383 |
| Coronary heart disease | 75,344 (41.8) | 60,396 (41.6) | 14,948 (42.8) | **<0.001** |
| Acute myocardial infarction | 3,891 (2.2) | 2,491 (1.7) | 1,400 (4.0) | **<0.001** |
| Prior myocardial infarction | 13,706 (7.6) | 10,680 (7.4) | 3,026 (8.7) | **<0.001** |
| Mitral valve disorder | 28,239 (15.7) | 22,857 (15.7) | 5,382 (15.4) | 0.135 |
| Aortic valve disorder | 18,270 (10.1) | 14,351 (9.9) | 3,919 (11.2) | **<0.001** |
| Dilated cardiomyopathy | 14,118 (7.8) | 11,521 (7.9) | 2,597 (7.4) | **0.002** |
| Prior ischemic stroke or ICB | 6,038 (3.4) | 4,605 (3.2) | 1,433 (4.1) | **<0.001** |
| Ventricular tachycardia | 2,301 (1.3) | 1,708 (1.2) | 593 (1.7) | **<0.001** |
| Renal failure | 91,920 (51.0) | 72,021 (49.5) | 19,899 (56.9) | **<0.001** |
| Diabetes | 70,469 (39.1) | 57,281 (39.4) | 13,188 (37.7) | **<0.001** |
| COPD | 31,517 (17.5) | 25,179 (17.3) | 6,338 (18.1) | **<0.001** |
| Pneumonia | 23,568 (13.1) | 17,491 (12.0) | 6,077 (17.4) | **<0.001** |
| Depression | 10,188 (5.7) | 7,657 (5.3) | 2,531 (7.2) | **<0.001** |
| Dementia | 15,920 (8.8) | 11,247 (7.7) | 4,673 (13.4) | **<0.001** |
| Solid tumor without metastasis | 3,410 (1.9) | 2,369 (1.6) | 1,041 (3.0) | **<0.001** |

<sup>1</sup>Pearson's Chi<sup>2</sup> test; Significant values are in bold; significance defined as P < .0025 after Bonferroni correction.
Abbreviations: No number; yrs years; NYHA New York Heart Association Functional Classification; ICB intracranial bleeding; COPD chronic obstructive pulmonary disease.

We found use of OAC in 80.6% (145,368) (Tab. 1) with a noticeable rise in prescription throughout the observation years (2017: 78.0%; 2018: 80.9%; 2019: 82.9%). Patient age and the degree of comorbidity differed between the OAC and non-OAC group. The median age for the OAC group was 81 years (IQR, 76 to 86 years) while patients belonging to the non-OAC group had a median age of 82 years (IQR, 77 to 88 years). The highest share of OAC patients were classified as NYHA III (42.8%) and for non-OAC patients as NYHA IV (44.0%). Comorbidities were significantly less present in patients treated with OACs, except for dilated cardiomyopathy (OAC: 7.9%; non-OAC: 7.4%) and diabetes (OAC: 39.4%; non-OAC: 37.7%) occurring more frequently in the OAC cohort and hypertension (OAC: 81.8%; non-OAC: 82.0%) as well as mitral valve disorder (OAC: 15.7%; non-OAC: 15.4%) being spread equally amongst both groups (Tab. 1).

Broken down by OAC type, VKAs were prescribed to 39,049 (21.7%) patients, FXals to 108.217 (60.0%) and DTIs to 6,189 (3.4%) patients, respectively (Supplementary (Supp.) Tab. 1). The distribution of sex amongst these subgroups varied with 57.7% of the FXaI cohort being female while in the DTI and VKA group, 54.8% and 50.0% were women. Throughout the observation years (2017/2018/2019), the use of VKAs (26.1%/ 21.3%/ 17.7%) and DTI (3.8%/ 3.5%/ 3.0%) dropped while FXaI prescriptions increased (52.8%/ 60.7%/ 66.4%). Overall, 8,087 patients had more than one prescription of an OAC type. In detail, 231 patients received VKAs and DTIs, 6,657 patients VKAs and FXaIs and 1,121 patients had a prescription of FXal and DTI. 39 patients were being prescribed with all three, VKA, DTI as well as FXal. The distribution of patients with and without an OAC combination therapy is visualized in form of a Venn diagram in Supp. Fig. 1.

### Mortality

A total of 33,531 (18.6%) patients died 91-365 days after discharge. In addition, 183 patients where censored as their AOK insurance ended prior to day 365. The Kaplan-Meier estimate for survival was 81.3% (95% CI 81.1% to 81.5%) with significant differences according to OAC (82.9% (95% CI 82.7% to 83.1%), compared to the non-OAC patient group (74.8% (95% CI 74.3% to 75.3%); log-rank test p <0.001). Details are provided in Fig. 2 (a). In the univariate analysis, significant differences in survival between each OAC group were noticeable with DTIs presenting the lowest rate (VKA (83.9% (95% CI 83.5% to 84.3%) vs. FXaI (84.6% (95% CI 83.6% to 85.6%) vs. DTI (82.5% (95% CI 82.3% to 82.7%)). The associated Kaplan-Meier curve is demonstrated in Fig. 2 (b).

**Figure 2.**
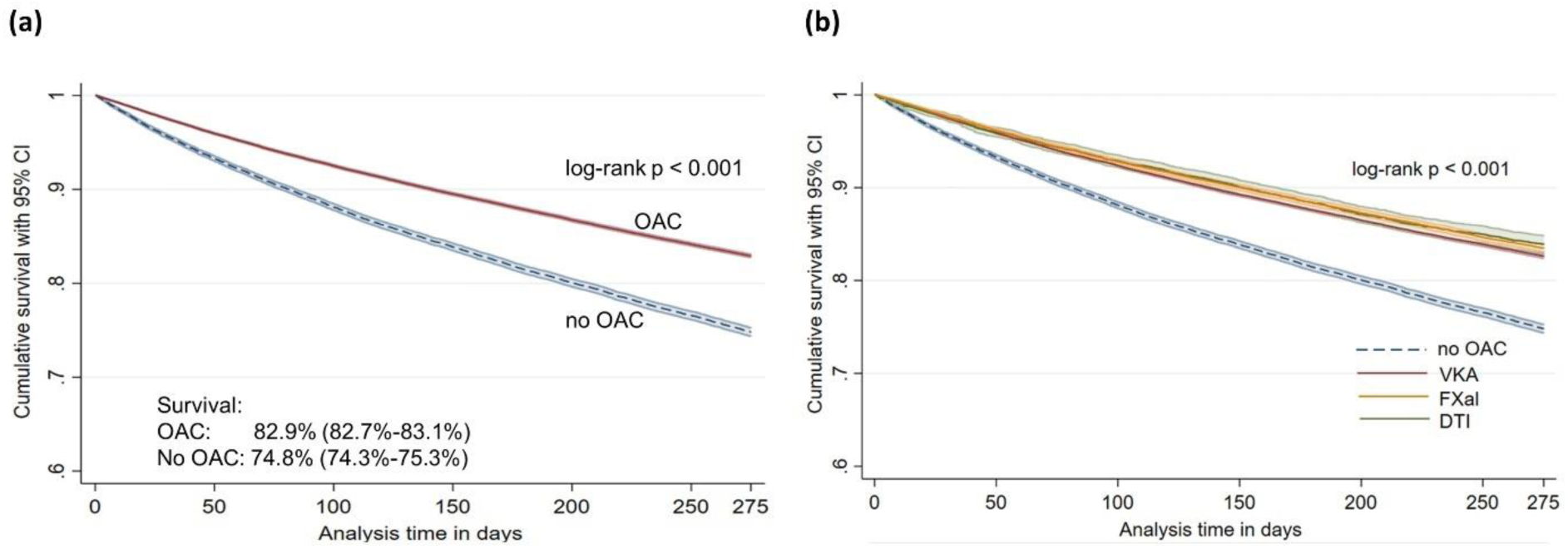
Kaplan-Meier survival according to (a) oral anticoagulation in general and (b) the specific type of oral anticoagulation. Kaplan-Meier curves representing the cumulative survival (with 95% confidence interval, CI) within 91-365 days after index hospitalization with a differentiation according to **(a)** the use of oral anticoagulants (OAC, red curve) versus the non-OAC patient group (blue curve) as well as according to **(b)** the specific types of OAC (red curve: VKA, orange curve: FXaI, green curve: DTI) versus the non-OAC group (blue curve); Log-rank test; significance defined as P < 0.05. Abbreviations: CI confidence interval; OAC oral anticoagulant; VKA vitamin K antagonist; FXaI factor Xa inhibitor; DTI direct-thrombin inhibitor.

Multivariable Cox proportional hazards survival analysis demonstrated that the risk of death was lower with the use of any OAC (HR 0.77, 95% CI 0.75 to 0.79). Beside the OAC therapy, female sex (HR 0.78, 95% CI 0.76 to 0.80) and obesity (HR 0.80, 95% CI 0.77 to 0.83) were associated with a lower mortality risk. Details of different risk factors on the hazard of survival are listed in Tab. 2. The various anticoagulants had no significantly different effect on survival (VKA: HR 0.73, 95% CI 0.71 to 0.76; FXaI: HR 0.77, 95% CI 0.75 to 0.78; DTI: HR 0.71, 95% CI 0.66 to 0.77).

**Table 2.**
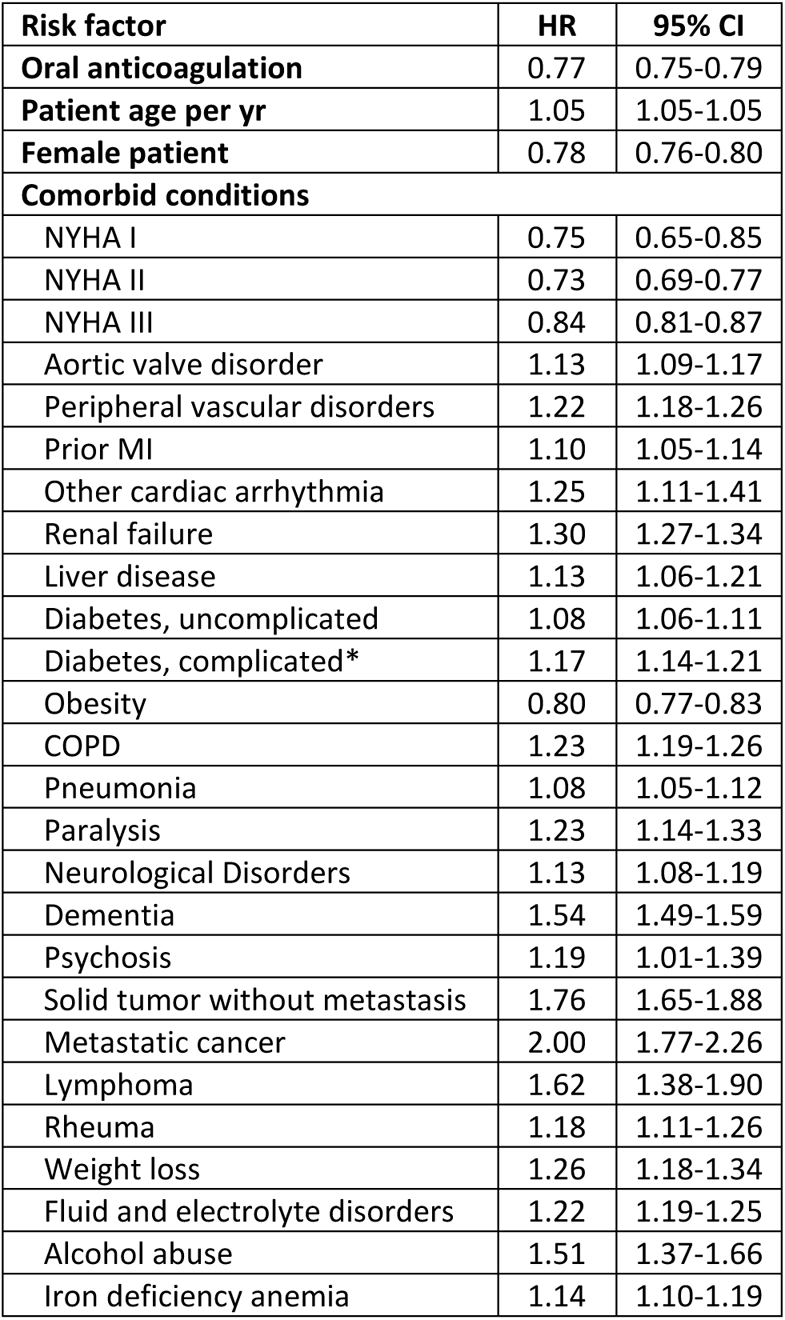
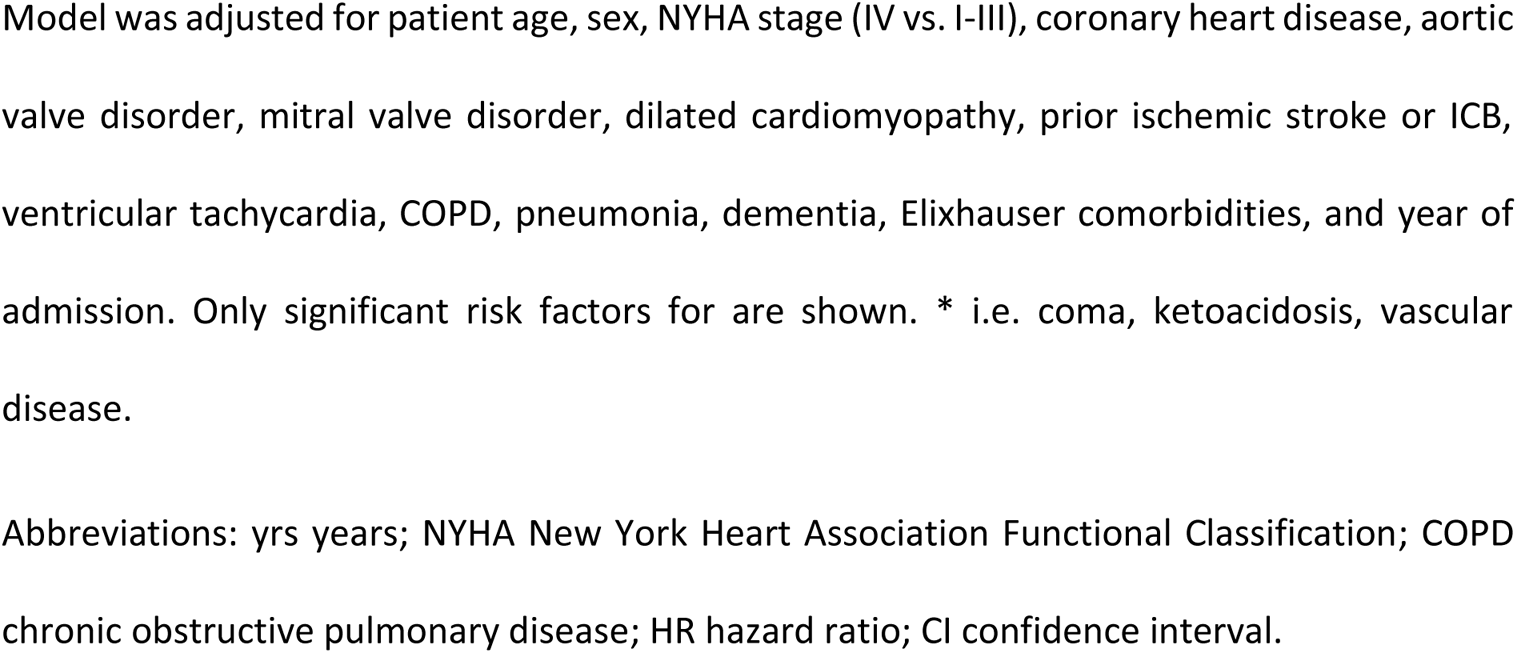
Effect of oral anticoagulation on hazard of survival within 91-365 days after admission.

| Risk factor | HR | 95% CI |
| --- | --- | --- |
| **Oral anticoagulation** | 0.77 | 0.75-0.79 |
| **Patient age per yr** | 1.05 | 1.05-1.05 |
| **Female patient** | 0.78 | 0.76-0.80 |
| **Comorbid conditions** |  |  |
| NYHA I | 0.75 | 0.65-0.85 |
| NYHA II | 0.73 | 0.69-0.77 |
| NYHA III | 0.84 | 0.81-0.87 |
| Aortic valve disorder | 1.13 | 1.09-1.17 |
| Peripheral vascular disorders | 1.22 | 1.18-1.26 |
| Prior MI | 1.10 | 1.05-1.14 |
| Other cardiac arrhythmia | 1.25 | 1.11-1.41 |
| Renal failure | 1.30 | 1.27-1.34 |
| Liver disease | 1.13 | 1.06-1.21 |
| Diabetes, uncomplicated | 1.08 | 1.06-1.11 |
| Diabetes, complicated* | 1.17 | 1.14-1.21 |
| Obesity | 0.80 | 0.77-0.83 |
| COPD | 1.23 | 1.19-1.26 |
| Pneumonia | 1.08 | 1.05-1.12 |
| Paralysis | 1.23 | 1.14-1.33 |
| Neurological Disorders | 1.13 | 1.08-1.19 |
| Dementia | 1.54 | 1.49-1.59 |
| Psychosis | 1.19 | 1.01-1.39 |
| Solid tumor without metastasis | 1.76 | 1.65-1.88 |
| Metastatic cancer | 2.00 | 1.77-2.26 |
| Lymphoma | 1.62 | 1.38-1.90 |
| Rheuma | 1.18 | 1.11-1.26 |
| Weight loss | 1.26 | 1.18-1.34 |
| Fluid and electrolyte disorders | 1.22 | 1.19-1.25 |
| Alcohol abuse | 1.51 | 1.37-1.66 |
| Iron deficiency anemia | 1.14 | 1.10-1.19 |

Model was adjusted for patient age, sex, NYHA stage (IV vs. I-III), coronary heart disease, aortic valve disorder, mitral valve disorder, dilated cardiomyopathy, prior ischemic stroke or ICB, ventricular tachycardia, COPD, pneumonia, dementia, Elixhauser comorbidities, and year of admission. Only significant risk factors for are shown. \* i.e. coma, ketoacidosis, vascular disease.
Abbreviations: yrs years; NYHA New York Heart Association Functional Classification; COPD chronic obstructive pulmonary disease; HR hazard ratio; CI confidence interval.

### Rehospitalization

Readmission for all causes within 91-365 days was at 29.9% (52,521/175,722) and occurred for ischemic stroke within 91-365 days in 1.9% of the cases (2,682/143,264). For these analyses 4,594, respectively 37,052 patients where censored prior to day 365 because of death or end of AOK insurance.

The Kaplan-Meier estimate for any cause readmission within 91-365 days was 70.9% (95% CI 70.7% to 71.1%) with significant differences according to OAC (with: 71.0%, 95% CI 70.8% to 71.3%; without: 70.2%, 95% CI 69.7% to 70.7%; log-rank test p <0.001). The same is accountable for the Kaplan-Meier estimate for ischemic stroke which was at 98.5% (95% CI 98.5% to 98.6%), likewise with significant differences according to the OAC use (with: 98.6% (95% CI 98.6% to 98.7%); without: 98.0% (95% CI 97.8% to 98.1%); log-rank test p <0.001). Both Kaplan-Meier survival curves are provided in Fig. 3.

**Figure 3.**
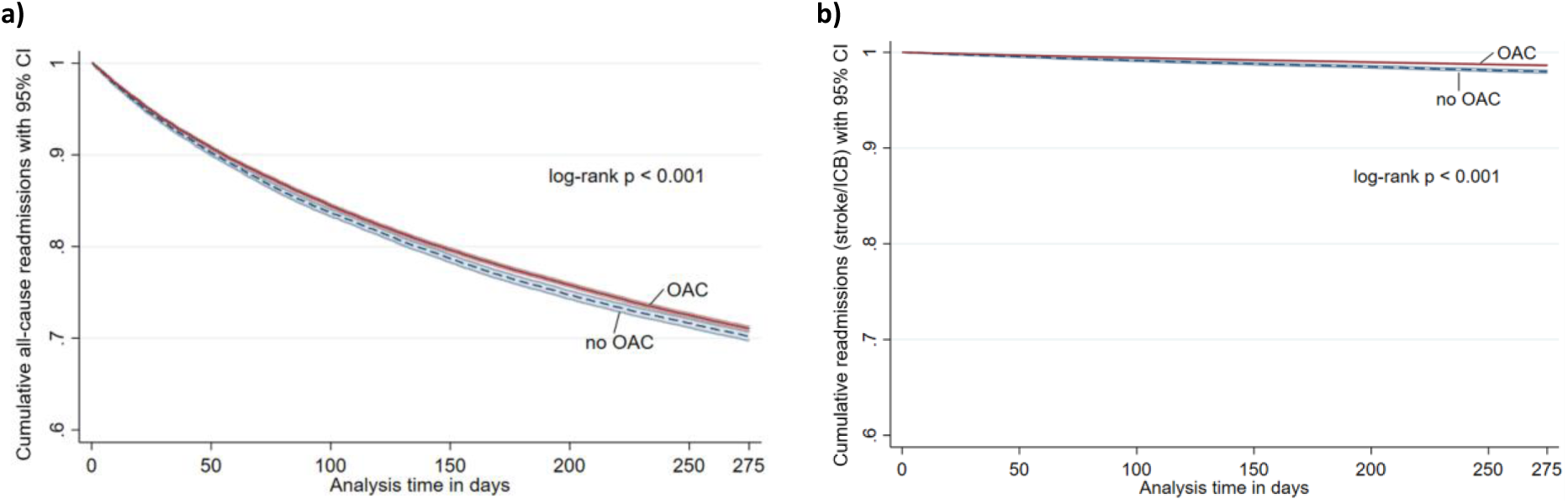
Kaplan Meier (a) all-cause readmissions and (b) rehospitalization due to ischemic stroke/ICB. Kapan-Meier curves representing **(a)** all-cause readmissions and **(b)** readmission due to ischemic stroke/ICB in the group of patients with oral anticoagulants (OAC, red curve) and non-OAC (blue curve); Log-rank test; significance defined as P < 0.05. Abbreviation: OAC oral anticoagulation.

Multivariable analysis demonstrated a significant benefit of OAC use on the total readmission rate (HR 0.97, 95% CI 0.94 to 0.99) (Tab. 3). Specifically, all-cause hospitalizations were the fewest in patients treated with FXaI (HR 0.96, 95% CI 0.93 to 0.98) in comparison to the VKA (HR 1.08, 95% CI 1.05 to 1.11) and DTI group (HR 1.02, 95% CI 0.96 to 1.08). A multivariable sub-analysis showed that readmission for stroke/ICB was likewise lower in patients with an OAC therapy (HR 0.71, 95% CI 0.65 to 0.77). The respective numbers are listed in Tab. 4. However, there were no differences visible, depending on the OAC type (VKA: HR 0.75, 95% CI 0.66 to 0.84); FXal: HR 0.69, 95% CI 0.63 to 0.75); DTI: HR 0.58, 95% CI 0.43 to 0.76).

**Table 3.**
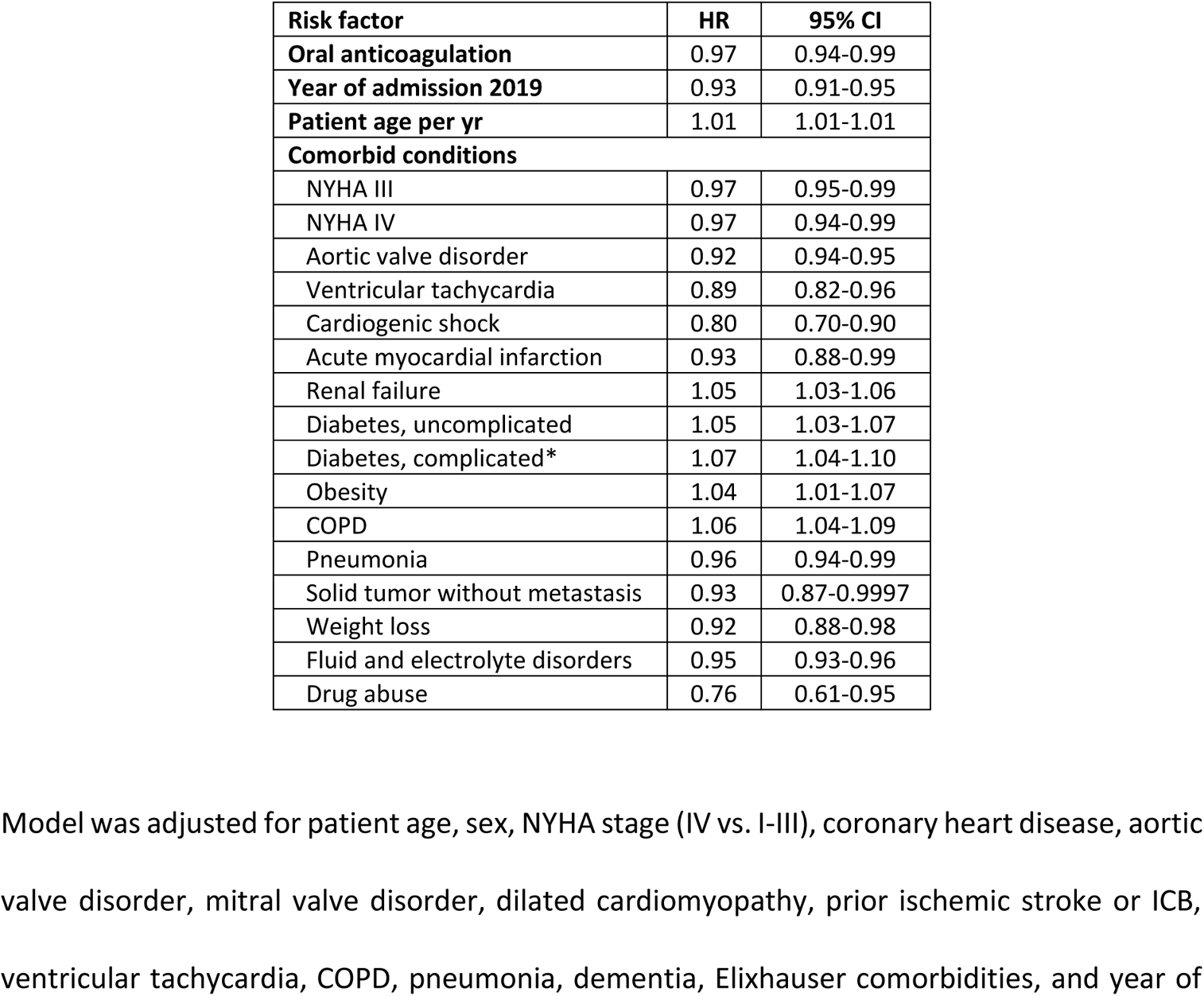
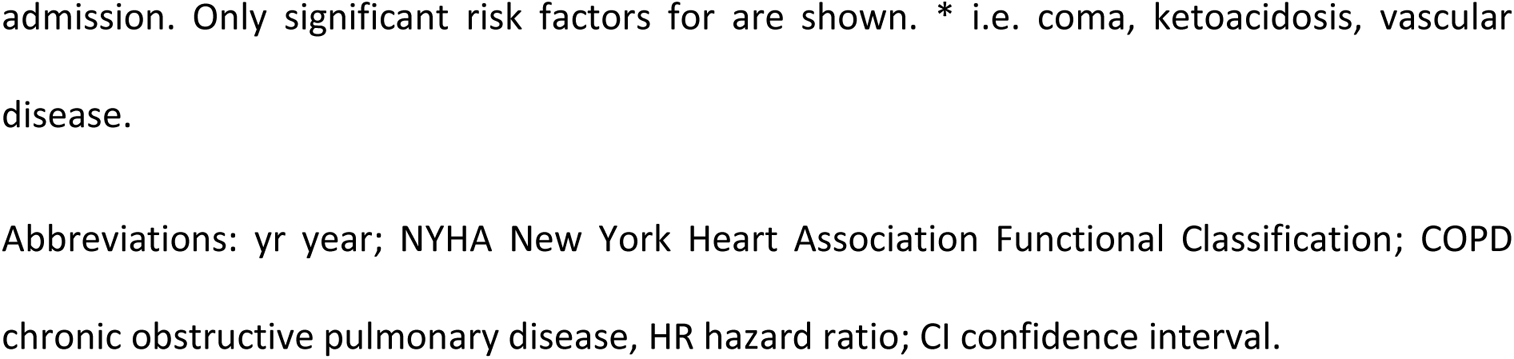
Effect of oral anticoagulation on hazard of all-cause readmission within 91-365 days after admission.

| Risk factor | HR | 95% CI |
| --- | --- | --- |
| Oral anticoagulation | 0.97 | 0.94-0.99 |
| Year of admission 2019 | 0.93 | 0.91-0.95 |
| Patient age per yr | 1.01 | 1.01-1.01 |
| **Comorbid conditions** |  |  |
| NYHA III | 0.97 | 0.95-0.99 |
| NYHA IV | 0.97 | 0.94-0.99 |
| Aortic valve disorder | 0.92 | 0.94-0.95 |
| Ventricular tachycardia | 0.89 | 0.82-0.96 |
| Cardiogenic shock | 0.80 | 0.70-0.90 |
| Acute myocardial infarction | 0.93 | 0.88-0.99 |
| Renal failure | 1.05 | 1.03-1.06 |
| Diabetes, uncomplicated | 1.05 | 1.03-1.07 |
| Diabetes, complicated* | 1.07 | 1.04-1.10 |
| Obesity | 1.04 | 1.01-1.07 |
| COPD | 1.06 | 1.04-1.09 |
| Pneumonia | 0.96 | 0.94-0.99 |
| Solid tumor without metastasis | 0.93 | 0.87-0.9997 |
| Weight loss | 0.92 | 0.88-0.98 |
| Fluid and electrolyte disorders | 0.95 | 0.93-0.96 |
| Drug abuse | 0.76 | 0.61-0.95 |

Model was adjusted for patient age, sex, NYHA stage (IV vs. I-III), coronary heart disease, aortic valve disorder, mitral valve disorder, dilated cardiomyopathy, prior ischemic stroke or ICB, ventricular tachycardia, COPD, pneumonia, dementia, Elixhauser comorbidities, and year of
admission. Only significant risk factors for are shown. \* i.e. coma, ketoacidosis, vascular disease.
Abbreviations: yr year; NYHA New York Heart Association Functional Classification; COPD chronic obstructive pulmonary disease, HR hazard ratio; CI confidence interval.

**Table 4.**
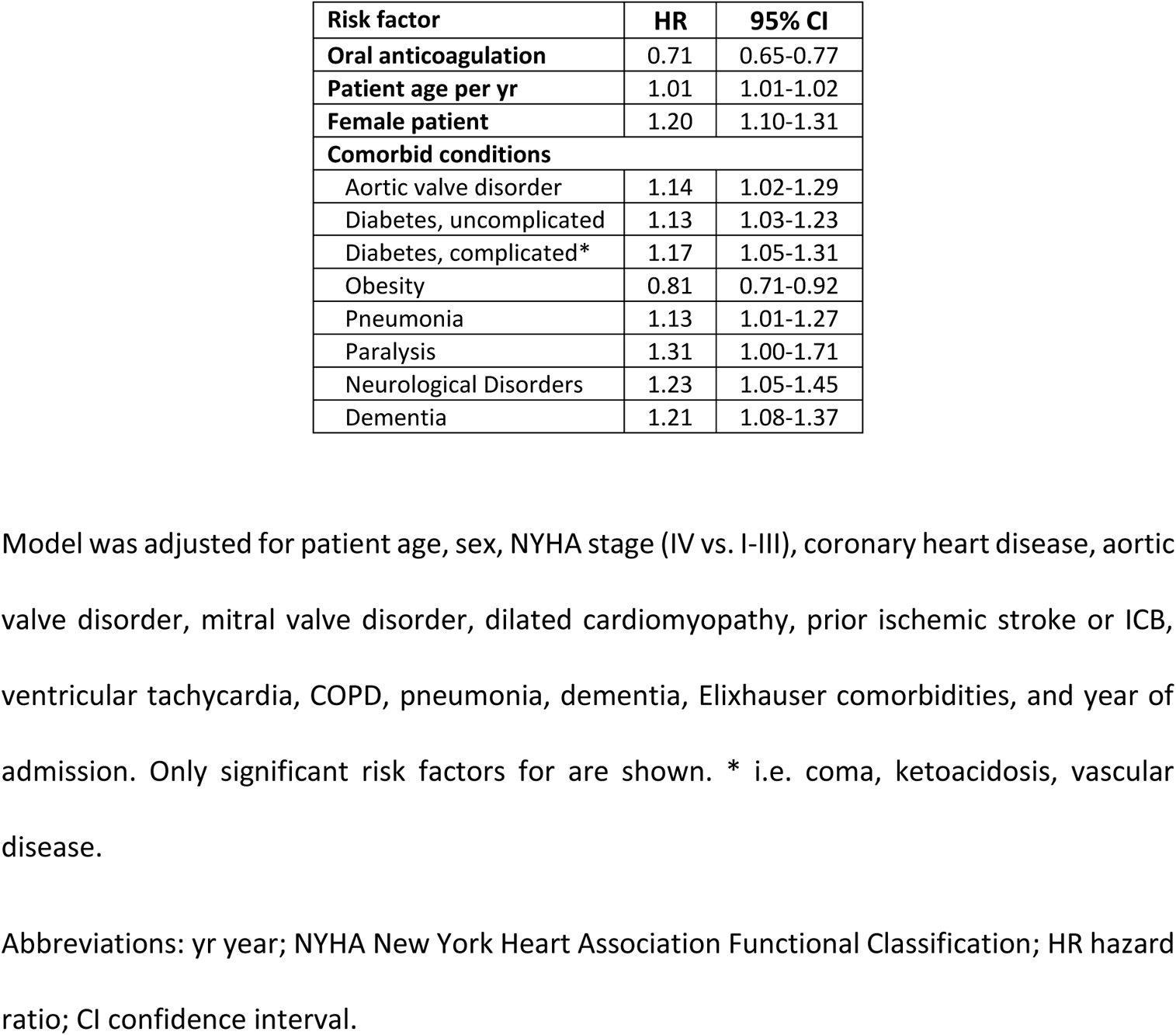
Effect of oral anticoagulation on hazard of readmission for stroke/ICB within 91-365 days after admission.

## Discussion

This routine data study analyzed one-year outcomes of OAC use in 180,316 patients suffering from HF complicated by AF between the years 2017-2019 and follow up until 2020. Firstly, it provides evidence that mortality between 91-365 days after index hospitalization reduces significantly in patients who are orally anticoagulated. Further, the secondary endpoint analysis of readmission rates showed a decrease in all-cause hospitalization as well as in readmissions due to complications of stroke and ICB after OAC use. Thirdly, by stratifying the OAC cohort, the use of VKA, FXaI and DTI was separately examined to evaluate additional benefits of NOACs. Here, prescription of FXaIs were associated with lower all-cause readmission, compared to other OAC subgroups, but no further NOAC superiority was registered.

Median age of HF-AF patients was slightly higher in our cohort (81 years) compared to other studies with a HF-AF subgroup analysis ^16, 18, 19, 25^. This might have resulted from the circumstance that the patients’ index hospital stay did not necessarily mark the HF onset but may have been caused by a decompensation phase due to disease progression. The comorbidity profile varies in existing literature with our cohort being roughly in line with it, while our 55.6% rate of females was comparatively higher ^18, 19, 25, 26^.

Our endpoint findings fully support the statements of currently existing ESC guidelines which recommend thromboembolic prophylaxis in HF-AF patients with OACs ^9^. OACs are mostly known for preventing thromboembolic complications like stroke ^27^. Accordingly, this effect was visible in our sample due to the significant decrease in stroke readmissions. However, beside this, rehospitalization and mortality rates were in total lower, indicating a broad beneficial effect of OACs on the progression of HF and AF that may not only work via anti-thromboembolism. Research so far has shown that OACs may also provide anti-inflammatory properties ^28–30^. Since inflammation is a key part of the pathomechanism behind HF as well as AF ^5, 8, 28, 29^, it can be assumed that OACs may also intervene in these processes and reduce mortality and morbidity in this way.

With regards to the overall OAC effect on the mortality trend, Lip et al. are not in line with our findings. They reported a continuously high one-year mortality in HF-AF patients although 82.1% of them were orally anticoagulated. Here, it must be kept in mind that the authors’ study setting differed as it was based on a survival analysis of HF versus non-HF as well as a stratification by HF subtype, limiting the comparability to our study ^22^. Generally, with a mortality risk of 18.6% in our HF-AF-OAC group (91-365 days after index hospital stay), the absolute rate remains high and may not reach the overall higher survival probability of AF patients without HF ^22^, despite OAC use. However, as visible in our data, OAC treatment may improve the fundamentally poor prognosis within the HF-AF cohort.

No major differences in effects were seen concerning the OAC type used; showing that NOACs are not clearly superior to VKAs when it comes to mortality, stroke/ICB as causes of inpatient stay in HF-AF. Only concerning the all-cause readmissions, FXaIs showed a comparatively better outcome than VKAs and DTIs. Therefore, our findings partly differed from existing trials that examined NOACs in a subgroup of AF-HF patients. In the ARISTOTLE trial, apixaban was described as more effective in reducing rates of stroke, systemic embolism, bleeding and mortality, being the only NOAC so far to show a survival benefit compared to warfarin ^19^. The RE-LY study revealed better outcome by the use of dabigatran (110mg) regarding major bleeding and, if a 150mg dosing regimen was applied, a reduction of stroke and systemic embolism were noticed ^18^. In accordance with our observations in the OAC group, Magnani et al. reported non-inferiority of edoxaban to warfarin ^17^ and van Diepen et al. demonstrated similar safety and efficacy of rivaroxaban and warfarin in AF-HF ^16^. As indicated by Xiong et al. in their AF-HF sub-analysis of these major trials, an explanatory approach for this inconsistency may lie in a dose-dependent variation of NOAC outcome ^20^; a factor that we did not evaluate. Simultaneously, it must be considered that these RCTs were primarily performed with warfarin as a representative of VKAs. However, in Germany, where our routine data origins from, the most commonly prescribed VKA is phenprocoumon - containing different pharmacological properties and so far, showing better outcomes than low-lose NOACs in AF patients ^31^. When interpreting our results, the non-inferiority of VKAs would implicate a certain flexibility in choosing the best fit for each HF-AF patient by also considering possible individual contraindications, comorbidities, HF subtype, and drug-drug interactions. This is highly relevant taking into account that more than 80% of HF patients have ≥ 2 comorbidities ^2^ and therefore may often experience polypharmacy ^32–34^. Beside these patient-related factors, the availability, medication costs and guidelines of different OACs vary globally ^34, 35^. In this context, OAC treatment for HF-AF patients can likewise be flexibly adapted to health care and physicians’ circumstances. Generally, we saw an increase in the prescription of FXaIs while VKA and DTI use reduced throughout the three observational years. These findings are in accordance with data on prescription trends, showing a down surge in VKA treatment and increasing FXaI prescriptions with simultaneous plateauing of DTI ^36, 37^.

The occurrence of HF-AF is often described as a dual epidemic in the literature ^38^ which demands for adequate approaches to slow down this development. Based on our data, an appropriate medical treatment with OACs in HF complicated by AF could provide a promising chance in reducing health care expenditures and burden on hospitals via decrease in readmission and mortality rate. Especially when considering the ageing population and improved diagnostics, both resulting in increasing disease prevalence ^2^, this finding appears crucial. To achieve this, patient adherence and constant re-evaluation by ambulatory doctors regarding the adequacy of the prescribed OAC is needed. As reported by Ozaki et al., real-world adherence to NOACs is yet not optimal although the absence of monitoring requirement in this medication group, contrasting to VKAs, would let us expect otherwise ^39^. Guidelines for patient education in medical treatment are provided by the ESC ^9^ which should be further implemented to rise adherence to OAC treatment, regardless of VKA or NOAC use. In prior studies, general follow-ups, including therapy re-evaluations, were already associated with better outcomes ^40^.

### Strengths and limitations

To our best knowledge, this routine data study is the first of its kind with a large, nationwide sample size providing valuable information on OAC therapy in HF patients with AF, supporting established guidelines. Based on the high patient number, a subgroup analysis of different OAC types was possible. Main limitations of this study are the retrospective methodology and possible inaccuracies in disease coding and extraction of medication data, which represents a frequent problem in routine data analysis. Also, since detailed clinical insights into the patients’ cases were not accessible, reasons for non-OAC use in HF-AF, despite an assumed indication, cannot be examined. The same accounts for patients who received more than one OAC. Here, a change in medication type (i.e. from VKA to NOAC) within the period from which pharmacy data was extracted, may explain the multiple prescriptions. Further on, only AOK patients were included, although other public as well as private insurances do exist in Germany. Nevertheless, it is the largest social health insurance ^23^, therefore AOK patients presumably represent the broad majority of German inhabitants across all social strata and milieus. This is highly relevant considering that the socioeconomic status and race are relevant influencing factor in HF and AF ^2, 41, 42^. Further, since ambulant prescription data was used to determine OAC medication, the outcome observation period was bound to the timing of quarterly billing and therefore started from 91 days after index hospitalization on. Also, no statements can be made about the exact time of OAC prescription. Here, it must be kept in mind that both mortality and stroke, are over proportionally high in the immediate period after hospital stay in patients with incident HF ^43^. Thus, the question of how prior or early OAC use influences mortality and rehospitalization in the immediate and critical time after discharge, remains unanswered and may be of high interest for future research.

### Conclusions

In conclusion, this routine data study on 180,316 patients shows a reduction in mortality, all-cause readmission, and readmission due to stroke/ICB in the observation period of 91-365 days after an index hospitalization for heart failure. These outcomes strongly underline the necessity of oral anticoagulation in HF-AF patients and mark their highly beneficial effect. Simultaneously, we recorded similar effects of VKAs and NOACs for these endpoints. For the clinical practice, this provides an opportunity to flexibly adapt the OAC medication to patients’ needs and circumstances. In the complex and heterogeneous group of HF-AF patients, advancing individualized treatment could thus represent one promising way to improve the disease prognosis.

## Data Availability

Data sharing is not applicable to this article as claim data were analyzed.

## Nonstandard Abbreviations and Acronyms

AF/Afib: atrial fibrillation
AOK: Allgemeine Ortskrankenkasse
HF: heart failure
ICB: intracerebral bleeding
ICD-10: international classification of diseases
NOAC: new oral anticoagulant
OAC: oral anticoagulant
VKA: vitamin K antagonist

## Acknowledgements

There are no acknowledgements to be mentioned.

